# Evidence for sexual antagonism and antagonistic pleiotropy in the maintenance of late onset Alzheimer’s disease alleles

**DOI:** 10.64898/2026.02.26.26347171

**Authors:** Edward H Morrow, Jon Alexander Harper

## Abstract

Trade-offs form a key constraint in many aspects of organismal evolution, though they may help maintain genetic diversity. Late-onset Alzheimer’s disease (LOAD) shows features in common with the male-female health survival paradox: females suffer from higher prevalence and risk, as well as faster rates of cognitive decline while males suffer higher mortality. Though antagonistic pleiotropy could explain the tendency of LOAD to appear late in life, the sexually dimorphic profile suggests a role for intralocus sexual conflict. Using published data on sex-specific genetic associations with LOAD risk, we found evidence for a number of sexually antagonistic loci, where alleles with net negative effects that reduce male risk but increase female risk are more common than alleles with reversed effects. Multiple lines of evidence also suggest there is an inverse relationship between cancer and LOAD risk. The combined effect of sexual antagonism and antagonistic pleiotropy could explain the persistence of alleles that increase LOAD risk in post-reproductive females, if they also reduce cancer risk in males. This framework could be applied to other female-biased late-life conditions, and our results may be useful in informing polygenic risk scores or therapies where genotype-by-sex effects may result in undesired outcomes for one particular sex.

## Background

Late onset Alzheimer’s disease (LOAD) is a multifactorial neurodegenerative disorder that causes a progressive decline in memory and cognition (Rabinovici, 2019). LOAD is increasing in prevalence and already affects tens of millions of people worldwide (*2024 Alzheimer’s disease facts and figures*, 2024). LOAD is the most common form of dementia and predominantly affects people aged 65 and over (*2024 Alzheimer’s disease facts and figures*, 2024), causing significant social and economic burdens globally, particularly among ageing populations (Skaria, 2022). As well as acting late in life, LOAD shows striking sexual dimorphism with higher prevalence, higher lifetime risk and faster rate of cognitive decline in women versus men (Fisher *et al*., 2018; Aggarwal & Mielke, 2023; Kolahchi *et al*., 2024), while men have higher mortality rates than women (Lapane *et al*., 2001; Sinforiani *et al*., 2010).

Several hypotheses have been proposed to explain the incidence and late-acting nature of LOAD (Fox, 2018). The most compelling is antagonistic pleiotropy, which is fundamentally a trade-off between reproduction and viability, where alleles that negatively affect viability or late-life health are maintained in a population if they also positively affect reproduction or early-life fitness components (Williams, 1957). However, antagonistic pleiotropy by itself cannot explain the sexually dimorphic profile of the disease. Instead the sex differences in prevalence and mortality of LOAD suggest intralocus sexual conflict (IASC) may play a role in maintaining some of the disease causing variants (Parker, 1979; Rice, 1992; Morrow & Connallon, 2013; Gilks *et al*., 2014). In this scenario, sexually antagonistic alleles are favoured by selection in one sex but are pathogenic when expressed in the opposite sex due to widespread sex-differences in biology. Theoretical models of balancing selection implicate both sexually antagonistic selection and antagonistic pleiotropy in the maintenance of genetic variation, either in isolation (Rice, 1984; Turelli & Barton, 2004) or together (Zajitschek & Connallon, 2018). Such models indicate that while the conditions for maintaining genetic variation under each form individually are quite narrow, the parameter space becomes broader when both forms act simultaneously.

The sex differences in risk profile, prevalence, progression and mortality of LOAD also reflect the so-called male-female health-survival paradox, which describes the generally observed phenomenon that women tend to live longer than men but in poorer health in later life (Oksuzyan *et al*., 2008). Interestingly, IASC has also been proposed as a resolution of this paradox (Archer *et al*., 2018), where male-benefit sexually antagonistic alleles with late-acting negative effects accumulate because their effects are not selected against in females due to reproductive menopause. From the perspective of LOAD, if late-acting LOAD alleles are in fact favoured by selection in males, then this IASC model could in principle explain why LOAD is female-biased. The post-reproductive period in females essentially generates a selection bias towards male-benefit late-acting alleles at the expense of female health. This framework, which takes into account the combined effects of antagonistic pleiotropy and IASC, could therefore improve our ability to explain the maintenance and evolutionary dynamics of LOAD causing alleles.

The genetic architecture of LOAD is highly polygenic and not yet fully resolved (Rosenthal & Kamboh, 2014; Ridge *et al*., 2016; Andrews *et al*., 2023; Bhardwaj *et al*., 2023). It is highly heritable, with a narrow-sense heritability of around 70% but with a substantial amount of ‘missing heritability’ (Andrews *et al*., 2023). The ancestral *ε4* allele of the Apolipoprotein E (*APOE*) gene is the major genetic risk factor for developing the disease (Bertram *et al*., 2007; Altmann *et al*., 2014; Dato *et al*., 2023). There is also a complex relationship between the vulnerability to LOAD that the *ε4* allele confers in women and decreasing oestrogen levels during menopause, generally by delaying menopause but exacerbating the negative effects of oestrogen loss (Gamache et al., 2020). The negative effects of the *ε4* allele on LOAD also extend to men and their late-life reductions in testosterone, but it is thought that since this decline in men is more gradual and occurs later in life the negative effects remain relatively greater in women (Gamache et al., 2020). *APOE* is a highly pleiotropic gene, with diverse effects on several other diseases and biological systems including cardiovascular disease, type II diabetes, as well as fertility and longevity. There is also evidence that the *ε4* allele is associated with lower female fertility than the *ε2* allele, but with greater longevity (Martínez-Martínez *et al*., 2020). Dato *et al*. (2023), have also found alleles at 8 loci (in 5 genes) show sexually antagonistic effects on LOAD risk when in epistasis with *APOE* alleles. Overall, at least 100 other loci have a significant genome-wide association with LOAD risk for individuals of European ancestry (Lambert *et al*., 2023), and reports of a further 18 loci show sex-dependent or sex-specific effects, some of which exhibit complex epistatic interactions with other loci (Gamache et al., 2020).

The evolutionary dynamics of alleles experiencing either IASC or antagonistic pleiotropy should depend on the relative magnitude of the effects in the two contexts. At sexually antagonistic loci, allele frequencies should correlate positively with their net effects across the two sexes such that alleles with net positive effects occur at higher equilibrium frequencies than alleles with net negative effects (Morrow & Connallon, 2013). This prediction was upheld in data from both flies and humans (Harper *et al*., 2021; Harper & Morrow, 2025). Similarly, allele frequencies at loci experiencing antagonistic pleiotropy should correlate with net effects on fitness across life-history traits. For example, if two alleles at a given locus with identical negative effects in late-life differed in the magnitude of early-life positive effects then we would expect the allele with the larger early-life effect to occur at the higher frequency. Under circumstances where a locus experiences both IASC and antagonistic pleiotropy, the predicted relationship between allele frequencies and their net effects across the sexes may hold for early-life fitness components but be reversed for late-life fitness components, when selection may be weaker. Importantly, the combined action of both antagonisms should facilitate the maintenance of genetic variation (Zajitschek & Connallon, 2018).

There is good evidence that both antagonistic pleiotropy and IASC are widespread evolutionary phenomena. The evolution of many fitness related traits is constrained by these mechanisms, either through the pleiotropic effects that genes have on multiple traits (Lemaître *et al*., 2015), or by sex-dependent and sexually antagonistic genetic effects (Ober *et al*., 2008; Gilks *et al*., 2014; Bernabeu *et al*., 2021; Harper *et al*., 2021). These genetic conflicts are also implicated across a broad range of human diseases (Carter & Nguyen, 2011; Byars & Voskarides, 2020), many of which show widespread sexual dimorphism (Mauvais-Jarvis *et al*., 2020). Given the ubiquity of antagonistic pleiotropy and the inevitability of sexual antagonism (Connallon & Clark, 2014), it is likely that both operate simultaneously for many traits (Wong & Holman, 2023). This possibility has received limited attention theoretically (Zajitschek & Connallon, 2018) and investigating this empirically is challenging since sets of genes experiencing both antagonistic pleiotropy and sexual antagonism must first be identified for fitness-related traits (Archer *et al*., 2018; Ruzicka *et al*., 2019, 2020; Jardine *et al*., 2021; Chakrabarty *et al*., 2024). However, evidence for a number of sexually antagonistic loci has recently been reported for LOAD (Prokopenko *et al*., 2020; see also Dato *et al*., 2023). We therefore take advantage of this recent evidence to investigate how the interplay between sexually antagonistic selection and antagonistic pleiotropy influence the evolutionary dynamics of alleles that underlie LOAD.

## Methods

We re-analysed data described in Prokopenko *et al*. (2020), which carried out sex-specific and sex stratified family-based association tests (FBATs) using data from four different sources. These included two different whole-genome sequencing studies: (i) the NIMH Alzheimer’s disease genetics initiative study (NIMH) (Blacker et al., 1997); and (ii) the family component of the National Institute of Aging Alzheimer’s Disease Sequencing Project (NIA-ADSP) sample (Beecham et al., 2017) - the NIMH and NIA-ADSP data were combined for their analysis. They also used two large genome-wide meta-analyses: (iii) Jansen *et al*. (455,000 individuals; 2019); and (iv) Kunkle *et al*. (90,000 individuals; 2019), which were analysed separately. Prokopenko *et al*. (2020) note that the sex-specific FBATs will have greater power to detect genetic effects in the opposite direction, a feature in common with other sex-differentiated tests (Magi et al., 2010). Conversely, the power to detect genetic effects that differ in magnitude but not direction will be lower. As a consequence, care must be taken when interpreting the relative abundances of sex-opposite and sex-specific or sex-limited effects.

From the combined analysis of the NIMH and NIA-ADSP data, Prokopenko *et al*. (2020) found 16 loci (in 9 different genes) that showed sex-specific associations and passed their stringent quality control (described in Prokopenko *et al*., 2020; see their Supplementary Table 1). From analyses of the two genome-wide meta-analyses, a further 29 loci (in 29 different genes) from Jansen *et al*. (2019), and 24 loci (in 24 different genes) from Kunkle *et al*. (2019) showed statistically significant sex-specific associations with AD risk (listed in Supplementary Tables 2 & 3, respectively in Prokopenko *et al*., 2020). There was little overlap across these 3 conservative analyses, with a total of 69 unique loci in 52 unique genes with sex-specific associations. We examined this combined list for loci with sexually antagonistic effects to form the first dataset for our analysis.

A less conservative fourth analysis by Prokopenko *et al*. (2020, their Supplementary Table 4), listed a large number of loci (n = 10,222) that were nominally significant for sex-specific associations with LOAD (P < 0.05 in genotype-by-sex interaction tests), but were 500kb up- or downstream of the variants reported in Jansen *et al*. (2019) and Kunkle *et al*. (2019). Loci with sexually antagonistic effects from this much larger list formed the second dataset for our analysis.

Sex-specific effect sizes from all tables (*z*-scores) were converted to Cohen’s *d* using the formulas in Koricheva *et al*. (2013). Male and female *d* values were calculated independently for each variant. Since positive effect sizes reported in Prokopenko *et al*. (2020) correspond to an increased risk of LOAD, we reversed the sign of all effect sizes to ensure deleterious effect sizes (in terms of LOAD risk) corresponded to negative values. Variants were filtered for a significant genotype-by-sex interaction (P < 0.05) and then filtered for loci that reported sexually antagonistic effects (*d* values in males and females with opposite signs). Sex-specific sample sizes were calculated directly from genotype counts when available (data originating from Prokopenko *et al*., 2020). When allele counts were unavailable (Jansen *et al*., 2019; Kunkle *et al*., 2019), male and female sample sizes were derived from the reported cohort-level sex proportions. Allele frequencies were then calculated from tables of effect and alternate allele counts. After filtering loci for sexually antagonistic effects we also limited the second group from the nominally significant loci to the top 100 loci ranked in reverse order of P-values.

We modelled how effect allele frequency varied with effect size ratio (the ratio of positive to negative effect sizes) using a generalized linear model (GLM). For sexually antagonistic loci, effect size ratios can take any negative value, but values between 0 and -1 indicate the balance of effects across the sexes is net negative, and values < -1 indicate net positive effects. The expectation was that higher sexually antagonistic allele frequencies would be associated with net beneficial effects when calculated across both sexes i.e. a negative relationship (Morrow & Connallon, 2013; Harper *et al*., 2021). The GLMs were fitted to both datasets with a quasibinomial error distribution with a logit link function to correct for overdispersion. Alleles were also coded into male-benefit/female-detriment or female-benefit/male-detriment groups based on the signs of the effect sizes. Differences in effect size ratios and effect allele frequencies between these two groups were tested with Mann-Whitney U tests with approximate P-values due to ties. All statistical analyses were performed in R (version 4.4.3). All data and code for the analysis and figures are publicly available (https://doi.org/10.5281/zenodo.18681253).

## Results

Alleles at all 16 loci from the NIMH and NIA-ADSP data, 14 loci from the Kunkle *et al*. (2019) data, and 17 loci from the Jansen *et al*. (2015) data had z-values with opposite signs from the male- or female-only FBAT analyses, indicating they were sexually antagonistic. However only 18 of these 47 loci (16 from the NIMH and NIA-ADSP data and 2 from Kunkle *et al*. (2019)), passed the filtering step of P ≤ 0.05 for the interaction test. These 18 formed the conservative set of sexually antagonistic loci for LOAD (Supplementary Table 1). Out of 10,222 loci with nominally significant associations with LOAD, 10,054 (98%) showed opposite effects in males and females (Supplementary Figure 1). From these we selected the top 100 for further analysis, based on lowest P-values as our second set of sexually antagonistic loci (equivalent to a P-value cutoff of ≤ 0.00773; Supplementary Table 2).

There was a significant positive relationship between effect size ratio and effect allele frequency for both the conservative set of loci (slope 2.46±0.399, P = 1.37x10^-5^, dispersion parameter 868, n = 18, Figure 1A) and the 100 nominally significant loci (slope 1.92±0.652, P = 3.97x10^-3^, dispersion parameter 191, n = 100, Figure 1B). There was also a clear distinction in the effect size ratios of male-benefit/female-detriment alleles and female-benefit/male-detriment alleles. In both sets, male-benefit alleles were dominated by net negative effects (90% and 100%, respectively) while female-benefit alleles were net positive on average (75% and 91%, respectively; upper two boxplots, Figure 1). The frequencies of male-benefit alleles were also higher in the top 100 nominally significant loci, with the smaller conservative set showing a similar but non-significant trend (two boxplots on right-hand sides, Figure 1). We also investigated whether these differences in net effects and allele frequencies between male- and female-benefit alleles were observed across all 10,054 nominally significant loci. Again, we found a clear pattern that male-benefit alleles have almost exclusively net negative effects on LOAD, while female-benefit alleles have net positive effects (Supplementary Figure 2).

**Figure 1.**
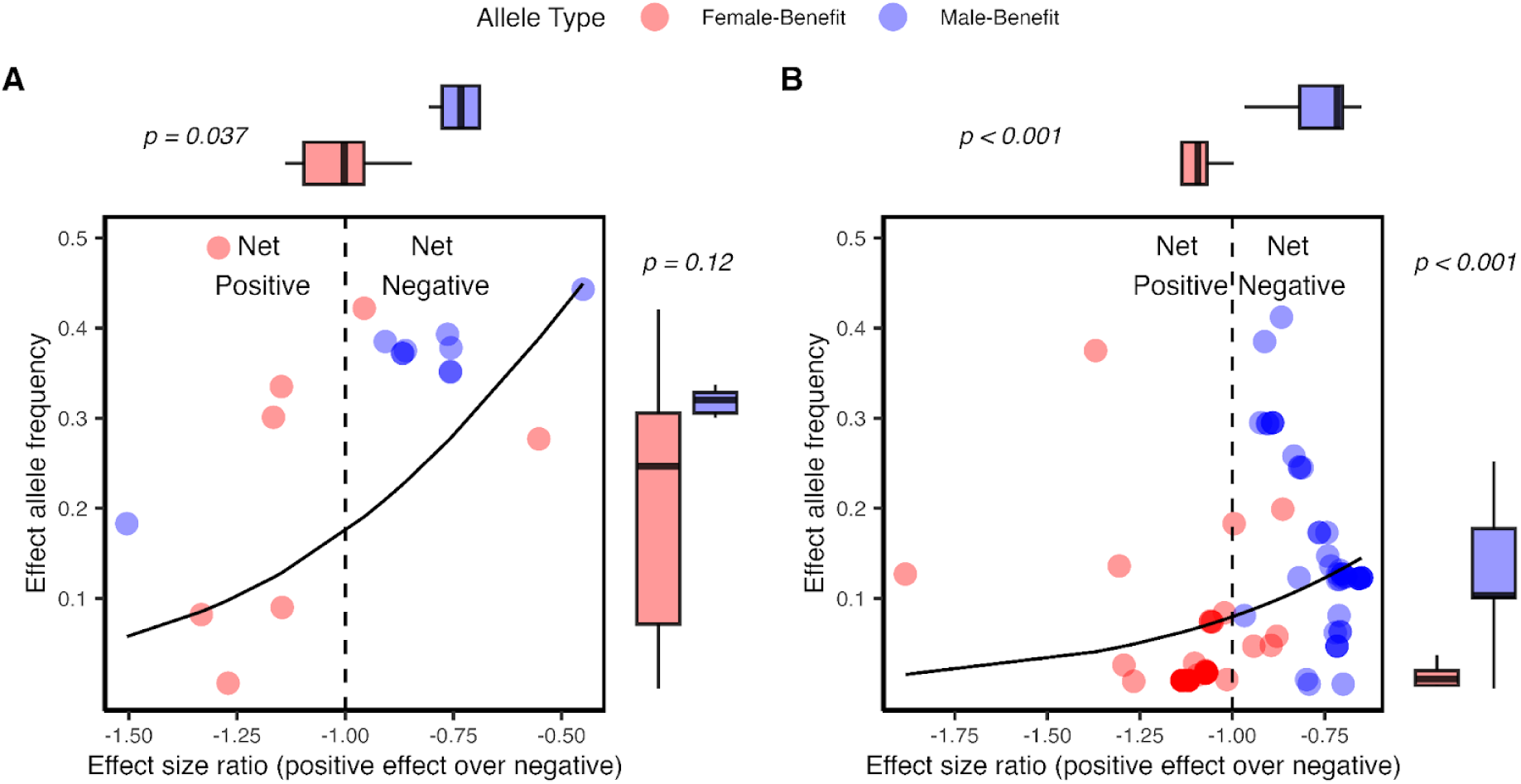
Effect allele frequency shows a positive relationship with effect size ratio for both A) the conservative set of loci and B) the nominally significant set of loci. Lines represent predicted relationships from the GLMs. Boxplots above panels compare effect size ratios between allele types (female-benefit and male-benefit alleles), while boxplots to the right of panels compare effect allele frequencies between allele types (P-values from Mann-Whitney U tests).

## Discussion

We investigated whether sexually antagonistic genes might explain the sexually dimorphic nature of LOAD, and resolve the apparent health-survival paradox for this important neurodegenerative disorder (Archer *et al*., 2018). Using data derived from four independent sources (Prokopenko *et al*., 2020) we found a large number of loci with sexually antagonistic effects, representing a significant increase in the total found in previous studies of sexual antagonism in human disease or quantitative traits (Winkler *et al*., 2015; Harper *et al*., 2021). Remarkably, alleles that increase LOAD risk in females but have male-benefit effects occur at higher frequencies and are more likely to have net negative effects than alleles that protect females from LOAD (Figure 1). The combined action of genetic variation at these loci could contribute in part to the sexual dimorphism widely observed in LOAD, since the most common sexually antagonistic alleles segregating in the population are those that harm females the most. Theory suggests that if antagonistic pleiotropy coincides with sexual antagonism, then genetic diversity may be more easily maintained (Zajitschek & Connallon, 2018). In support of this we find male-benefit alleles in both sets are in some cases as common as the most common LOAD associated loci (Andrews *et al*., 2023). LOAD therefore represents a unique case of a human disease that not only shows significant differences between the sexes in its phenotypic expression but also a clear sexually antagonistic component in its genetic architecture. Effects at these loci would therefore account for some of the missing heritability observed for LOAD (Karlsson *et al*., 2022), although they warrant further validation in additional datasets.

The result that sexually antagonistic alleles with net negative effects on LOAD occur at higher frequencies than alleles with net positive effects is both counterintuitive and opposite to the pattern previously found in sets of sexually antagonistic alleles associated with human disease (Harper *et al*., 2021), and fitness in flies (Harper & Morrow, 2025). However, this result can be understood if there is a trade-off between LOAD risk and another fitness component or disease. A primary candidate is cancer, as there is substantial evidence for antagonistic pleiotropy between LOAD and several tumour types (Bassil et al., 2024), along with some mechanistic details for how these two diseases are inversely linked. For example, data from the Framingham Heart Study, while controlling for survivor bias, found that the protective effect of one disease over the other was considerable: cancer survivors had a 33% reduction in risk for Alzheimer’s and Alzheimer’s patients had a 61% decreased risk of cancer (Driver et al., 2012). Other studies (Roe et al., 2005, 2010; Musicco et al., 2013) and a recent metanalysis (Zheng *et al*., 2025) report effects of similar magnitude. Evidence from therapeutic interventions also point to a pleiotopic and mechanistic link between the two, where cancer treatment with Tamoxifen or chemotherapy reduces LOAD risk (Ma et al., 2025). Remarkably, transplantation of tumour cells to healthy mouse models of Alzheimer’s disease reduced amyloid pathology and rescued their cognition (Li *et al*., 2026). This effect is mediated through the action of the protein cystatin-C, which plays a role in immune system regulation and is secreted by some cancer cells. In the mouse brain, cystatin-C activates TREM2 that targets for destruction the toxic amyloid plaques that are present in LOAD (Li et al., 2026). Thus, cystatin-C may in part be responsible for the antagonistic pleiotropy observed between LOAD and cancer.

An inverse relationship between LOAD and cancer suggests that sexually antagonistic alleles that increase female LOAD risk could be favoured by selection in males via reductions in cancer risk. Since LOAD acts during late/post-reproductive life of females it may escape selection, while cancer would reduce male fitness at any age. As a result the frequency of male-benefit/female-detriment alleles may be higher than alleles that protect females but harm males (Figure 2). If common, genetic variation at these loci would generate a negative cross-trait cross-sex genetic correlation between LOAD and cancer. A testable prediction then is that the alleles that modify male and female LOAD risk in opposite directions should also modify cancer risk in similar sex-dependent ways. The complementary genetic correlation between male LOAD risk and female cancer (not shown) may be more balanced in terms of selection pressures, since both LOAD in males and cancer in females would lower fitness. An exception to this is if LOAD risk loci impacted cancer risk in male-limited tissues, such as the reproductive system. Then the risk for females would be selectively neutral, and selection would again favour male-benefit alleles (reducing their LOAD risk).

**Figure 2.**
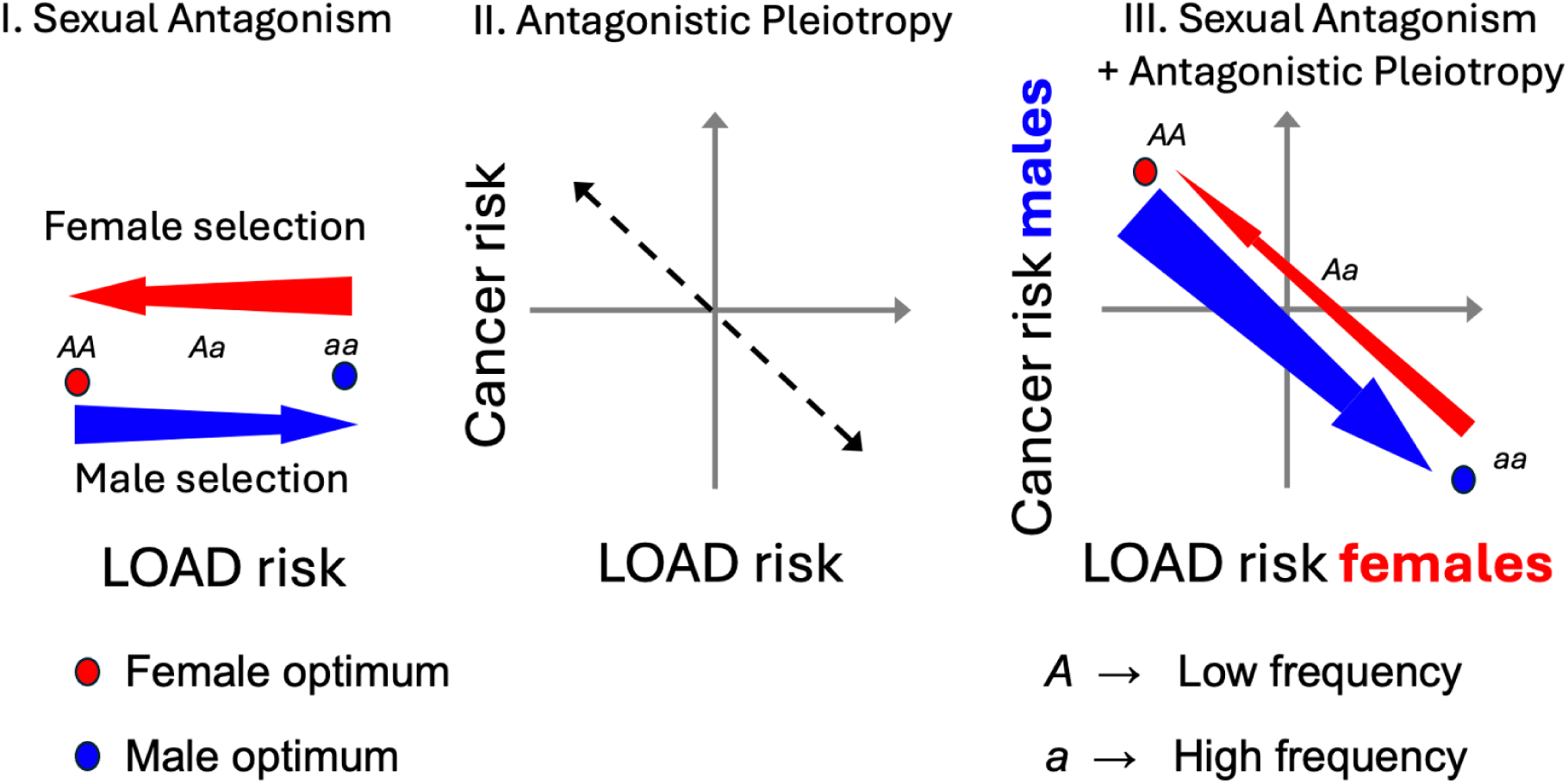
**I.** Under sexual antagonism only, and considering LOAD risk in isolation: Allele *A* is female-benefit/male-detriment allele and *a* is male-benefit/female detriment allele, as they move the two sexes closest to their respective optima. Each additional *a* allele increases risk for LOAD in females but reduces it for males. **II.** Under antagonistic pleiotropy only, but considering both diseases: Substitution of alleles that increase LOAD risk reduces cancer risk, and vice versa, generating an inverse relationship between both diseases. **III.** Combining both sexual antagonism and antagonistic pleiotropy: Looking at a cross-trait cross-sex genetic correlation, each additional *a* allele in females increases LOAD risk but has the reverse effect on cancer risk in males. Since there will be stronger selection against cancer in males (thick blue arrow) than the selection against LOAD in females (thin red arrow), *a* alleles are expected to be at a higher frequency than *A* alleles.

Additional mechanistic links between LOAD and cancer have been proposed, including the general view that both diseases are connected by the common processes of cell survival/cell death (Zabłocka *et al*., 2021). One specific mechanism involves the signalling protein Pin1, which catalyses the *cis*-to-*trans* isomerisation of Amyloid Precursor Protein (APP) and tau, changing the conformations of proteins from a pathogenic form (*cis*) to a healthy one (*trans*). Low Pin1 expression is associated with LOAD and a polymorphism in Pin1 that increase its expression (rs2287839) is associated with later LOAD age-of-onset, while a polymorphism resulting in lower Pin1 expression (rs2233678) is associated with decreased risk of different cancers (Driver & Lu, 2010; Lanni *et al*., 2021). This inverse Cancer-Alzheimer correlation through PIN1 hypothesis is further supported by a large cross-sectional analysis where cancer types that had low Pin1 expression had higher likelihood of secondary diagnosis of LOAD than cancer types with high expression of Pin1 (Sherzai et al., 2020).

Another candidate is the protease ADAM10, which cleaves APP, releasing a neuroprotective fragment called sAPPα and thereby reducing the production of the toxic amyloid-beta protein (Alexandre-Silva & Cominetti, 2024). ADAM10 activity is decreased in LOAD patients and also *in vitro* by the *APOE*-*ε4* allele (Shackleton *et al*., 2017; Alexandre-Silva & Cominetti, 2024). Conversely, ADAM10 is overexpressed in several different cancers, where it promotes cell proliferation and metastasis (Alexandre-Silva & Cominetti, 2024). Signalling pathways such as Wnt or the tumour suppressor p53 are also involved and interlinked with Pin1, which regulates both p53 and Wnt, while ADAM10 is intrinsically linked to all three (Alexandre-Silva & Cominetti, 2024). p53 activity by itself also increases LOAD risk (Zabłocka *et al*., 2021). Other potential molecular mechanisms include microRNAs, some of which show inverse patterns of regulation/activity between LOAD and cancer (Zabłocka *et al*., 2021).

Inflammation is a signature of both diseases and it is thought that dysregulation of the immune system can result in either cancer on one hand, if the immune system is not active enough in eliminating cancer cells, or LOAD on the other, if there is chronic neuroinflammation (Zabłocka *et al*., 2021). Supporting this hypothesis there is some evidence that treatment of cancer patients with immunosuppressives, reduces their risk of developing LOAD and some other neurocognitive disorders (Akushevich *et al*., 2021).

Several specific cancers show an inverse relationship with LOAD. These include prostate, breast, lung, melanoma and colorectal cancer (Bassil et al., 2024). Sherzai et al. (2020) found all 10 cancer types investigated showed the inverse relationship LOAD, with prostate, ovarian, and lung cancers displaying the strongest patterns. Ma et al. (2025) found lower LOAD risk for patients with colorectal, non-melanoma skin cancers and lung cancer, while LOAD patients had lower risk of lung, bladder, breast and colorectal cancers. Another longitudinal study found a particularly strong reduction in LOAD risk (hazard ratio = 0.53) for male genital cancers (Zhang *et al*., 2022). There is also evidence that while the *APOE*-*ε2* allele is protective and *ε4* is a risk factor for LOAD, the risk of these alleles is reversed for melanoma/skin cancer (Ostendorf et al., 2020; Adaku et al., 2023).

Prostate cancer is particularly interesting from the perspective of the male-female health-survival paradox, as correct prostate functioning is essential for male fertility, and will experience natural (and possibly sexual) selection throughout life. It is also of course a male-limited tissue, which means there is no source of selection via females. Prostate cancer is inversely related to cognitive performance and dementia; both are thought to be mediated by chronic inflammation and testosterone titre (Mekli et al., 2023; Oseni et al., 2023). Androgen Deprivation Therapy is used to reduce the stimulating effect that testosterone has on tumour cell growth during early stages of prostate cancer. However, it has been found that reducing testosterone levels increases the risk of developing various types of dementia (Motlagh *et al*., 2021; Ma *et al*., 2025), as testosterone is neuroprotective (Ketchem et al., 2023). The androgen receptor gene (*AR*) also shows antagonistic pleiotropy between cancer and fertility in both sexes (Carter & Nguyen, 2011). There is also some limited evidence for differences between *APOE* genotypes and male fertility (Gerdes *et al*., 1996; Corbo *et al*., 2004; Setarehbadi *et al*., 2012). Overall, it is possible that the positive effects of male-benefit sexually antagonistic alleles are mediated via selection on the male reproductive system, protecting it from cancer and promoting fertility, though many other cancer types shared between the sexes are clearly implicated in the inverse relationship with LOAD.

## Conclusions

Multiple lines of evidence indicate that some of the polygenic architecture of LOAD involves sexually antagonistic loci, and that allele frequencies at those loci are highest when alleles are beneficial in terms of male LOAD, at the expense of females, even when the net effects across both sexes are negative. We hypothesize that the well-supported inverse relationship between LOAD and cancer may help explain this counterintuitive result, if alleles that increase female LOAD risk post-reproduction also reduce cancer risk in males during earlier life-stages. The effect of male-benefit alleles may be particularly large if they enhance male fertility and/or protect the male reproductive tract from cancer. Clearly, there is scope for sexually antagonistic genetic effects in advancing our understanding of the genetic basis of both diseases, and may be worthwhile to include them in calculations of polygenic risk scores. This view is further supported by evidence of epistatic interactions between the major effect locus *APOE* and sexually antagonistic loci (Dato *et al*., 2023), and that *APOE* alleles exhibit a pleiotropic relationship with cancer (Ostendorf *et al*., 2020; Adaku *et al*., 2023).

There may be other examples of female-biased late-life diseases that are driven by trade-offs with other traits/diseases and sexual conflict, including stroke (Rexrode *et al*., 2022), thyroid disease (Mulder, 1998), osteoporosis (Alswat, 2017), and some autoimmune diseases, such as rheumatoid arthritis (Gerosa *et al*., 2008). In those cases, a first step would be to perform sex-differentiated GWAS, which have sufficient power to detect effects with opposite signs in the two sexes (Magi *et al*., 2010). Finally, the inverse relationship between cancer and LOAD may also have consequences for therapeutic design, where targeting specific proteins (e.g. ADAM10) to treat cancer, may result in unintended and undesirable consequences for the risk profile of LOAD (Alexandre-Silva & Cominetti, 2024). Our analysis indicates that the direction of those effects may also be genotype- and sex-dependent. One example for this problem has already been noted, where doses of intranasal insulin in *APOE*-*ε4*-negative men with LOAD showed cognitive improvement, while *APOE*-*ε4*-negative women showed further cognitive decline (Claxton *et al*., 2013). In contrast, *APOE*-*ε4*-positive individuals showed no effects of the intervention.

## Data Availability

All data produced are available online at: https://doi.org/10.5281/zenodo.18681253

## Acknowledgements

We thank the evolution group at Karlstad University for helpful discussions and Matthew Neale for providing funding to JAH.

## Author contributions

EHM: Conceptualization; Formal analysis; Funding acquisition; Investigation; Methodology; Project administration; Supervision; Validation; Visualization; Writing – original draft; Writing – review & editing. JAH: Data curation; Formal analysis; Investigation; Methodology; Validation; Visualization; Writing – review & editing.

## Declaration of AI use

We have used AI-assisted technologies for trouble-shooting some summary and plotting parts of the R code.

## Supplementary Material

**Supplementary Figure 1.**
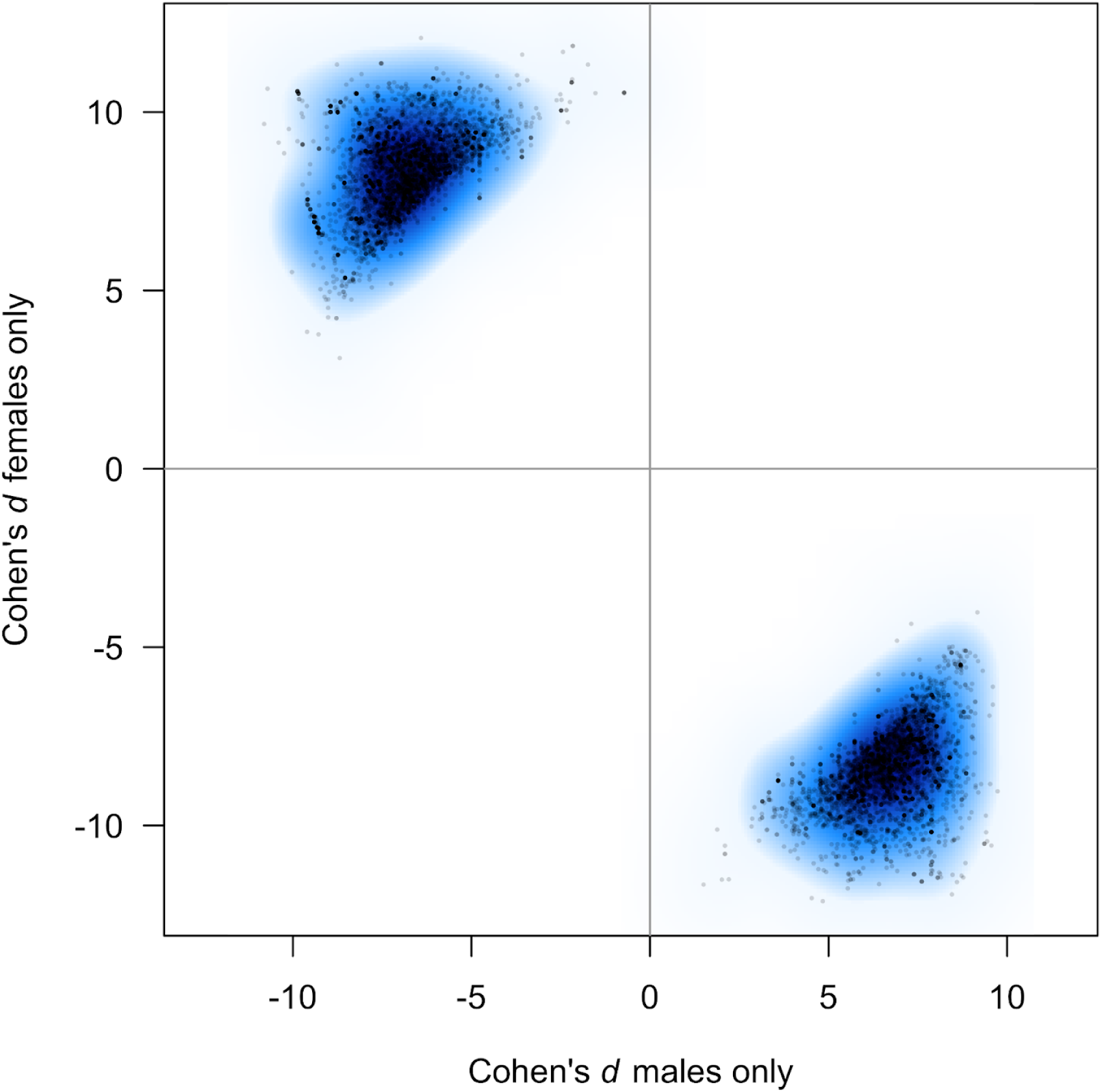
Distribution of male and female effects from the nominally significant set of loci (n = 10,222) that show sex-specific associations with LOAD (P < 0.05 in genotype-by-sex interaction tests), 500kb up- or downstream of the variants reported in Jansen *et al*. (2019) and Kunkle *et al*. (2019).

**Supplementary Figure 2.**
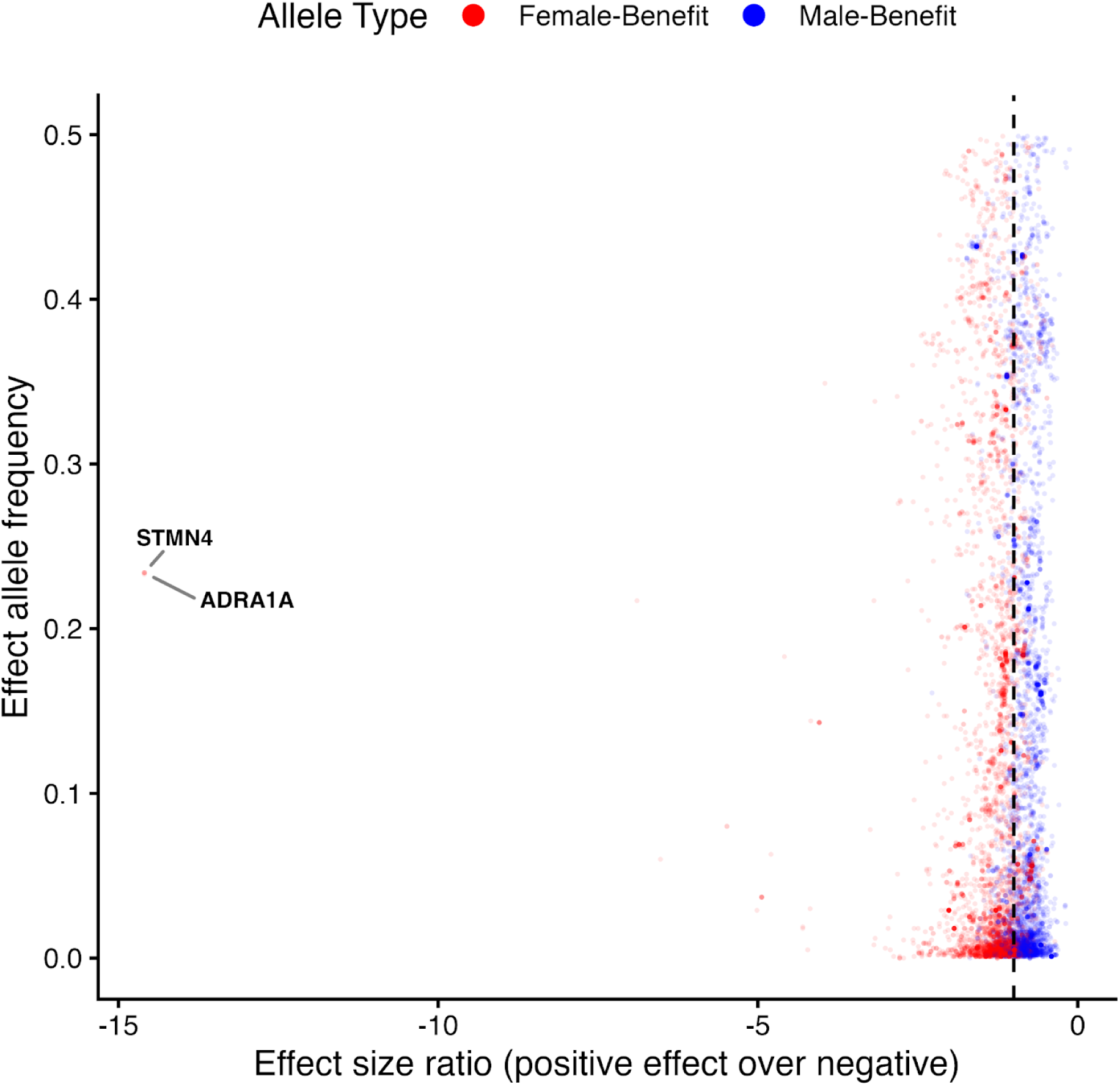
Effect size ratio and effect allele frequency for the set of nominally significant sexually antagonistic loci (n = 10,054). As in the main Figure 1, the vertical broken line at effect size ratio -1 is the boundary between net positive effects (to the left of the line) and net negative effects (to the right of the line). Effect size ratio differed significantly between male- and female-benefit alleles (Mann-Whitney U test, P < 2.2 x 10^-16^), but effect allele frequency did not (P = 0.129). Five loci from 2 genes were outliers with effect size ratios < -14 (labelled on figure).

## Supplementary Tables

[Supplementary_Table_1](https://docs.google.com/spreadsheets/d/17mBoU_clEL0NEsyzPzAq5kzyv52jaCTkmDX7ieyOFgI/edit?usp=sharing)

[Supplementary_Table_2](https://docs.google.com/spreadsheets/d/1VhqE8tKVzWoKXjNUqWtKVDD8XmkpbCaGwjr0a87oywM/edit?usp=sharing)

## References

2024 Alzheimer’s disease facts and figures. 2024. Alzheimer’s Association.

Adaku, N., Ostendorf, B.N., Mei, W. & Tavazoie, S.F. 2023. Apolipoprotein E2 Stimulates Protein Synthesis and Promotes Melanoma Progression and Metastasis. Cancer Res. 83: 3013–3025.

Aggarwal, N.T. & Mielke, M.M. 2023. Sex Differences in Alzheimer’s Disease. Neurol. Clin. 41: 343–358.

Akushevich, I., Yashkin, A.P., Kravchenko, J. & Kertai, M.D. 2021. Chemotherapy and the Risk of Alzheimer’s Disease in Colorectal Cancer Survivors: Evidence From the Medicare System. JCO Oncol. Pract. 17: e1649–e1659.

Alexandre-Silva, V. & Cominetti, M.R. 2024. Unraveling the dual role of ADAM10: Bridging the gap between cancer and Alzheimer’s disease. Mech. Ageing Dev. 219: 111928.

Alswat, K.A. 2017. Gender Disparities in Osteoporosis. J. Clin. Med. Res. 9: 382–387.

Altmann, A., Tian, L., Henderson, V.W., Greicius, M.D. & Investigators, A.D.N.I. 2014. Sex modifies the APOE-related risk of developing Alzheimer disease. Ann. Neurol. 75: 563–573.

Andrews, S.J., Renton, A.E., Fulton-Howard, B., Podlesny-Drabiniok, A., Marcora, E. & Goate, A.M. 2023. The complex genetic architecture of Alzheimer’s disease: novel insights and future directions. eBioMedicine 90. Elsevier.

Archer, C.R., Recker, M., Duffy, E. & Hosken, D.J. 2018. Intralocus sexual conflict can resolve the male-female health-survival paradox. Nat. Commun. 9: 5048. Nature Publishing Group.

Bassil, D.T., Zheng, B., Su, B., Kafetsouli, D., Udeh-Momoh, C., Tzoulaki, I., et al. 2024. Lower Incidence of Dementia Following Cancer Diagnoses: Evidence from a Large Cohort and Mendelian Randomization Study. J. Prev. Alzheimers Dis. 11: 1397–1405.

Beecham, G.W., Bis, J.C., Martin, E.R., Choi, S.-H., DeStefano, A.L., van Duijn, C.M., et al. 2017. The Alzheimer’s Disease Sequencing Project: Study design and sample selection. Neurol. Genet. 3: e194. Wolters Kluwer.

Bernabeu, E., Canela-Xandri, O., Rawlik, K., Talenti, A., Prendergast, J. & Tenesa, A. 2021. Sex differences in genetic architecture in the UK Biobank. Nat. Genet. 53: 1283–1289.

Bertram, L., McQueen, M.B., Mullin, K., Blacker, D. & Tanzi, R.E. 2007. Systematic meta-analyses of Alzheimer disease genetic association studies: the AlzGene database. Nat. Genet. 39: 17–23. Nature Publishing Group.

Bhardwaj, A., Liyanage, S.I. & Weaver, D.F. 2023. Cancer and Alzheimer’s Inverse Correlation: an Immunogenetic Analysis. Mol. Neurobiol. 60: 3086–3099.

Blacker, D., Haines, J.L., Rodes, L., Terwedow, H., Go, R.C.P., Harrell, L.E., et al. 1997. ApoE-4 and Age at Onset of Alzheimer’s Disease. Neurology 48: 139–147. Wolters Kluwer.

Byars, S.G. & Voskarides, K. 2020. Antagonistic Pleiotropy in Human Disease. J. Mol. Evol. 88: 12–25. Springer US.

Carter, A.J. & Nguyen, A.Q. 2011. Antagonistic pleiotropy as a widespread mechanism for the maintenance of polymorphic disease alleles. BMC Med. Genet. 12: 160.

Chakrabarty, A., Chakraborty, S., Nandi, D. & Basu, A. 2024. Multivariate genetic architecture reveals testosterone-driven sexual antagonism in contemporary humans. Proc. Natl. Acad. Sci. U. S. A. 121: e2404364121.

Claxton, A., Baker, L.D., Wilkinson, C.W., Trittschuh, E.H., Chapmana, D., Watson, G.S., et al. 2013. Sex and ApoE Genotype Differences in Treatment Response to Two Doses of Intranasal Insulin in Adults with Mild Cognitive Impairment or Alzheimer’s Disease. J. Alzheimers Dis. JAD 35: 789–797.

Connallon, T. & Clark, A.G. 2014. Evolutionary inevitability of sexual antagonism. Proc. Biol. Sci. 281: 20132123.

Corbo, R.M., Scacchi, R. & Cresta, M. 2004. Differential reproductive efficiency associated with common apolipoprotein E alleles in postreproductive-aged subjects. Fertil. Steril. 81: 104–107.

Dato, S., De Rango, F., Crocco, P., Pallotti, S., Belloy, M.E., Le Guen, Y., et al. 2023. Sex- and APOE-specific genetic risk factors for late-onset Alzheimer’s disease: Evidence from gene–gene interaction of longevity-related loci. Aging Cell 22: e13938.

Driver, J.A., Beiser, A., Au, R., Kreger, B.E., Splansky, G.L., Kurth, T., et al. 2012. Inverse association between cancer and Alzheimer’s disease: results from the Framingham Heart Study. BMJ 344: e1442. British Medical Journal Publishing Group.

Driver, J.A. & Lu, K.P. 2010. Pin1: a new genetic link between Alzheimer’s disease, cancer and aging. Curr. Aging Sci. 3: 158–165.

Fisher, D.W., Bennett, D.A. & Dong, H. 2018. Sexual dimorphism in predisposition to Alzheimer’s disease. Neurobiol. Aging 70: 308–324.

Fox, M. 2018. ‘Evolutionary medicine’ perspectives on Alzheimer’s Disease: Review and new directions. Ageing Res. Rev. 47: 140–148. Elsevier.

Gamache, J., Yun, Y. & Chiba-Falek, O. 2020. Sex-dependent effect of APOE on Alzheimer’s disease and other age-related neurodegenerative disorders. Dis. Model. Mech. 13: dmm045211.

Gerdes, L.U., Gerdes, C., Hansen, P.S., Klausen, I.C. & Færgeman, O. 1996. Are men carrying the apolipoprotein ε4- or ε2 allele less fertile than ε3ε3 genotypes? Hum. Genet. 98: 239–242.

Gerosa, M., De Angelis, V., Riboldi, P. & Meroni, P. 2008. Rheumatoid Arthritis: A Female Challenge. Womens Health 4: 195–201. SAGE Publications Ltd STM.

Gilks, W.P., Abbott, J.K. & Morrow, E.H. 2014. Sex differences in disease genetics: evidence, evolution, and detection. Trends Genet. 30: 453–463.

Harper, J.A., Janicke, T. & Morrow, E.H. 2021. Systematic review reveals multiple sexually antagonistic polymorphisms affecting human disease and complex traits. Evolution 75: 3087–3097.

Harper, J.A. & Morrow, E.H. 2025. The adaptive value of recombination in resolving intralocus sexual conflict by gene duplication. Proc. R. Soc. B Biol. Sci. 292: 20242629. Royal Society.

Jansen, I.E., Savage, J.E., Watanabe, K., Bryois, J., Williams, D.M., Steinberg, S., et al. 2019. Genome-wide meta-analysis identifies new loci and functional pathways influencing Alzheimer’s disease risk. Nat. Genet. 51: 404–413. Nature Publishing Group.

Jardine, M.D., Ruzicka, F., Diffley, C., Fowler, K. & Reuter, M. 2021. A non-coding indel polymorphism in the fruitless gene of Drosophila melanogaster exhibits antagonistically pleiotropic fitness effects. Proc. R. Soc. B Biol. Sci. 288: 20202958. Royal Society.

Karlsson, I.K., Escott-Price, V., Gatz, M., Hardy, J., Pedersen, N.L., Shoai, M., et al. 2022. Measuring heritable contributions to Alzheimer’s disease: polygenic risk score analysis with twins. Brain Commun. 4. Oxford Academic.

Ketchem, J.M., Bowman, E.J. & Isales, C.M. 2023. Male sex hormones, aging, and inflammation. Biogerontology 24: 1–25.

Kolahchi, Z., Henkel, N., Eladawi, M.A., Villarreal, E.C., Kandimalla, P., Lundh, A., et al. 2024. Sex and Gender Differences in Alzheimer’s Disease: Genetic, Hormonal, and Inflammation Impacts. Int. J. Mol. Sci. 25: 8485.

Koricheva, J., Gurevitch, J. & Mengersen, K. (eds). 2013. Handbook of Meta-analysis in Ecology and Evolution. Princeton University Press.

Kunkle, B.W., Grenier-Boley, B., Sims, R., Bis, J.C., Damotte, V., Naj, A.C., et al. 2019. Genetic meta-analysis of diagnosed Alzheimer’s disease identifies new risk loci and implicates Aβ, tau, immunity and lipid processing. Nat. Genet. 51: 414–430. Nature Publishing Group.

Lambert, J.-C., Ramirez, A., Grenier-Boley, B. & Bellenguez, C. 2023. Step by step: towards a better understanding of the genetic architecture of Alzheimer’s disease. Mol. Psychiatry 28: 2716–2727.

Lanni, C., Masi, M., Racchi, M. & Govoni, S. 2021. Cancer and Alzheimer’s disease inverse relationship: an age-associated diverging derailment of shared pathways. Mol. Psychiatry 26: 280–295. Nature Publishing Group.

Lapane, K.L., Gambassi, G., Landi, F., Sgadari, A., Mor, V. & Bernabei, R. 2001. Gender differences in predictors of mortality in nursing home residents with AD. Neurology 56: 650–654. Wolters Kluwer.

Lemaître, J.-F., Berger, V., Bonenfant, C., Douhard, M., Gamelon, M., Plard, F., et al. 2015. Early-late life trade-offs and the evolution of ageing in the wild. Proc. R. Soc. B Biol. Sci. 282: 20150209.

Li, X., Tang, X., Zeng, J., Duan, L., Hou, Z., Li, L., et al. 2026. Peripheral cancer attenuates amyloid pathology in Alzheimer’s disease via cystatin-c activation of TREM2. Cell, doi: 10.1016/j.cell.2025.12.020.

Ma, L., Tan, E.C.K., Goudey, B., Jin, L. & Pan, Y. 2025. Unraveling the bidirectional link between cancer and dementia and the impact of cancer therapies on dementia risk: A systematic review and meta-analysis. Alzheimers Dement. 21: e14540.

Magi, R., Lindgren, C.M. & Morris, A.P. 2010. Meta-analysis of sex-specific genome-wide association studies. Genet. Epidemiol. 34: 846–853.

Martínez-Martínez, A.B., Torres-Perez, E., Devanney, N., Del Moral, R., Johnson, L.A. & Arbones-Mainar, J.M. 2020. Beyond the CNS: The many peripheral roles of APOE. Neurobiol. Dis. 138: 104809.

Mauvais-Jarvis, F., Bairey Merz, N., Barnes, P.J., Brinton, R.D., Carrero, J.-J., DeMeo, D.L., et al. 2020. Sex and gender: modifiers of health, disease, and medicine. Lancet Lond. Engl. 396: 565–582.

Mekli, K., Lophatananon, A., Maharani, A., Nazroo, J.Y. & Muir, K.R. 2023. Association between an inflammatory biomarker score and future dementia diagnosis in the population-based UK Biobank cohort of 500,000 people. PLOS ONE 18: e0288045. Public Library of Science.

Morrow, E.H. & Connallon, T. 2013. Implications of sex-specific selection for the genetic basis of disease. Evol. Appl. 6: 1208–1217.

Motlagh, R.S., Quhal, F., Mori, K., Miura, N., Aydh, A., Laukhtina, E., et al. 2021. The Risk of New Onset Dementia and/or Alzheimer Disease among Patients with Prostate Cancer Treated with Androgen Deprivation Therapy: A Systematic Review and Meta-Analysis. J. Urol., doi: 10.1097/JU.0000000000001341. Wolters KluwerPhiladelphia, PA.

Mulder, J.E. 1998. THYROID DISEASE IN WOMEN. Med. Clin. North Am. 82: 103–125.

Musicco, M., Adorni, F., Di Santo, S., Prinelli, F., Pettenati, C., Caltagirone, C., et al. 2013. Inverse occurrence of cancer and Alzheimer disease. Neurology 81: 322–328. Wolters Kluwer.

Ober, C., Loisel, D.A. & Gilad, Y. 2008. Sex-specific genetic architecture of human disease. Nat Rev Genet 9: 911–922.

Oksuzyan, A., Juel, K., Vaupel, J.W. & Christensen, K. 2008. Men: good health and high mortality. Sex differences in health and aging. Aging Clin. Exp. Res. 20: 91–102.

Oseni, S.O., Naar, C., Pavlović, M., Asghar, W., Hartmann, J.X., Fields, G.B., et al. 2023. The Molecular Basis and Clinical Consequences of Chronic Inflammation in Prostatic Diseases: Prostatitis, Benign Prostatic Hyperplasia, and Prostate Cancer. Cancers 15: 3110. Multidisciplinary Digital Publishing Institute.

Ostendorf, B.N., Bilanovic, J., Adaku, N., Tafreshian, K.N., Tavora, B., Vaughan, R.D., et al. 2020. Common germline variants of the human APOE gene modulate melanoma progression and survival. Nat. Med. 26: 1048–1053.

Parker, G.A. 1979. Sexual selection and sexual conflict. In: Sexual selection and reproductive competition in insects, pp. 123–166. Academic Press, London.

Prokopenko, D., Hecker, J., Kirchner, R., Chapman, B.A., Hoffman, O., Mullin, K., et al. 2020. Identification of Novel Alzheimer’s Disease Loci Using Sex-Specific Family-Based Association Analysis of Whole-Genome Sequence Data. Sci. Rep. 10: 5029. Nature Publishing Group.

Rabinovici, G.D. 2019. Late-onset Alzheimer Disease. Contin. Lifelong Learn. Neurol. 25: 14–33.

Rexrode, K.M., Madsen, T.E., Yu, A.Y.X., Carcel, C., Lichtman, J.H. & Miller, E.C. 2022. The Impact of Sex and Gender on Stroke. Circ. Res. 130: 512–528. American Heart Association.

Rice, W.R. 1984. Sex chromosomes and the evolution of sexual dimorphism. Evolution 38: 735–742.

Rice, W.R. 1992. Sexually antagonistic genes - experimental evidence. Science 256: 1436–1439.

Ridge, P.G., Hoyt, K.B., Boehme, K., Mukherjee, S., Crane, P.K., Haines, J.L., et al. 2016. Assessment of the genetic variance of late-onset Alzheimer’s disease. Neurobiol. Aging 41: 200.e13–200.e20.

Roe, C.M., Behrens, M.I., Xiong, C., Miller, J.P. & Morris, J.C. 2005. Alzheimer disease and cancer. Neurology 64: 895–898. Wolters Kluwer.

Roe, C.M., Fitzpatrick, A.L., Xiong, C., Sieh, W., Kuller, L., Miller, J.P., et al. 2010. Cancer linked to Alzheimer disease but not vascular dementia. Neurology 74: 106–112. Wolters Kluwer.

Rosenthal, S.L. & Kamboh, M.I. 2014. Late-Onset Alzheimer’s Disease Genes and the Potentially Implicated Pathways. Curr. Genet. Med. Rep. 2: 85–101.

Ruzicka, F., Dutoit, L., Czuppon, P., Jordan, C.Y., Li, X.-Y., Olito, C., et al. 2020. The search for sexually antagonistic genes: Practical insights from studies of local adaptation and statistical genomics. Evol. Lett. 4–5: 398–415.

Ruzicka, F., Hill, M.S., Pennell, T.M., Flis, I., Ingleby, F.C., Mott, R., et al. 2019. Genome-wide sexually antagonistic variants reveal long-standing constraints on sexual dimorphism in fruit flies. PLoS Biol. 17: e3000244.

Setarehbadi, R., Vatannejad, A., Vaisi-Raygani, A., Amiri, I., Esfahani, M., Fattahi, A., et al. 2012. Apolipoprotein E genotypes of fertile and infertile men. Syst. Biol. Reprod. Med. 58: 263–267. Taylor & Francis.

Shackleton, B., Crawford, F. & Bachmeier, C. 2017. Apolipoprotein E-mediated Modulation of ADAM10 in Alzheimer’s Disease. Curr. Alzheimer Res. 14: 578–585. Bentham Science Publishers.

Sherzai, A.Z., Parasram, M., Haider, J.M. & Sherzai, D. 2020. Alzheimer Disease and Cancer: A National Inpatient Sample Analysis. Alzheimer Dis. Assoc. Disord. 34: 122.

Sinforiani, E., Citterio, A., Zucchella, C., Bono, G., Corbetta, S., Merlo, P., et al. 2010. Impact of Gender Differences on the Outcome of Alzheimer’s Disease. Dement. Geriatr. Cogn. Disord. 30: 147–154. S. Karger AG.

Skaria, A.P. 2022. The economic and societal burden of Alzheimer disease: managed care considerations. Am. J. Manag. Care 28: S188–S196.

Turelli, M. & Barton, N.H. 2004. Polygenic variation maintained by balancing selection: Pleiotropy, sex-dependent allelic effects and GxE interactions. Genetics 166: 1053–1079.

Williams, G.C. 1957. Pleiotropy, Natural Selection, and the Evolution of Senescence. Evolution 11: 398–411. [Society for the Study of Evolution, Wiley].

Winkler, T.W., Justice, A.E., Graff, M., Barata, L., Feitosa, M.F., Chu, S., et al. 2015. The Influence of Age and Sex on Genetic Associations with Adult Body Size and Shape: A Large-Scale Genome-Wide Interaction Study. PLOS Genet. 11: e1005378.

Wong, H.W.S. & Holman, L. 2023. Pleiotropic fitness effects across sexes and ages in the Drosophila genome and transcriptome. Evolution 77: 2642–2655.

Zabłocka, A., Kazana, W., Sochocka, M., Stańczykiewicz, B., Janusz, M., Leszek, J., et al. 2021. Inverse Correlation Between Alzheimer’s Disease and Cancer: Short Overview. Mol. Neurobiol. 58: 6335–6349.

Zajitschek, F. & Connallon, T. 2018. Antagonistic pleiotropy in species with separate sexes, and the maintenance of genetic variation in life-history traits and fitness. Evolution 72: 1306–1316.

Zhang, D.-D., Ou, Y.-N., Yang, L., Ma, Y.-H., Tan, L., Feng, J.-F., et al. 2022. Investigating the association between cancer and dementia risk: a longitudinal cohort study. Alzheimers Res. Ther. 14: 146.

Zheng, G., Xu, M., Dong, Z., Abdelrahman, Z. & Wang, X. 2025. Meta-analysis reveals an inverse relationship between Alzheimer’s disease and cancer. Behav. Brain Res. 478: 115327.

